# Predictors of symptomatic laboratory-confirmed SARS-COV-2 reinfection

**DOI:** 10.1101/2020.09.28.20203190

**Authors:** Efrén Murillo-Zamora, Carlos M. Hernandez-Suarez, Agustin Lugo-Radillo, Felipe Aguilar-Sollano, Oliver Mendoza-Cano

## Abstract

**Objective:** To identify factors predicting symptomatic laboratory-positive SARS-COV-2 reinfection.

**Method:** We conducted a nationwide retrospective cohort study and data from 99,993 confirmed cases of COVID-19 were analyzed.

**Results:** The overall risk of reinfection (28 or more elapsed days between both episodes onset) was 0.21%, and older subjects and those with mild primary disease were at reduced risk of reinfection. Healthcare workers and immunosuppressed or renal patients had at greater risk of SARS-COV-2 reinfection.

**Conclusions:** If replicated in other populations, these results may be useful to prioritize efforts focusing on the reduction of SARS-COV-2 spread and the related burden.

## Introduction

The COVID-19 (coronavirus disease 2019) by SARS-COV-2 (severe acute respiratory coronavirus 2) pandemic is a complex phenomenon and reinfection is one of the many ongoing related debates [1]. Current knowledge regarding factors predicting the SARS-COV-2 reinfection risk is scarce, and it has significant implications in public health policies, including vaccination strategies and relaxation of social distancing measures [2].

The social and economic burden of the COVID-19 pandemic in Mexico has been high, and by September 24, more than 715 thousand confirmed cases and nearly 80 thousand deaths had been registered [3]. This study aimed to evaluate factors predicting SARS-COV-2 symptomatic reinfection in a large and nationwide cohort of laboratory-confirmed COVID-19 survivors.

## Methods

We performed a nationwide retrospective cohort study in Mexico, including adults (aged 20 years or above) with laboratory-confirmed COVID-19 (quantitative reverse transcription-polymerase chain reaction, RT-qPCR) COVID-19 by SARS-COV-2. This analysis took place in September 2020, and a broader description of the methods has already been published [4]. Adults whose symptoms appeared from March to June 2020 and who recovered to primary infection, were analyzed. The primary binary outcome was symptomatic reinfection of SARS-COV-2 and was defined by the reappearance of symptoms of COVID-19 at 28 days or more after initial illness [5] and a positive RT-qPCR result. Risk ratios (RR) and 95% confidence intervals (CI), calculated using linear regression models, were used to identify factors associated with the risk of reinfection.

## Results

Data from 99,993 participants were analyzed and the overall risk of SARS-COV-2 symptomatic reinfection was 0.21% (*n* = 210). An RT-qPCR result between both episodes was available for 14 reinfection-positive subjects and all of them were negative.

The mean elapsed days (± standard deviation) between COVID-19 episodes were 61.0 ± 31.0 and ranged from 28 to 116. Mild subsequent illness was documented in 169 patients (80.5%) of reinfected subjects and the observed fatality rate was 4.3%.

Table 1 shows the characteristics of the study sample according to the reinfection status for selected variables. Patients with SARS-COV-2 reinfection were younger and were more likely to be healthcare professionals or other related employments. They were also more likely to have had milder symptoms at primary disease. They had a significantly higher prevalence of chronic kidney disease or immunosuppression (any cause except type 2 diabetes mellitus o kidney disease).

**Table 1.** Characteristics of the study sample according to symptomatic SARS-COV-2 reinfection status, Mexico 2020

|  | Overall |  | SARS-COV-2 reinfection |  |  |  |  |
| --- | --- | --- | --- | --- | --- | --- | --- |
|  |  |  | No |  | Yes |  | *p* |
|  | *n* = 99,993 |  | *n* = 99,783 |  | *n* = 210 |  |  |
| **Gender** |  |  |  |  |  |  |  |
| Female | 50,916 | (50.9) | 50,805 | (50.9) | 111 | (52.9) | 0.574 |
| Male | 49,077 | (49.1) | 48,978 | (49.1) | 99 | (47.1) |  |
| **Age (mean ± SD, years)** | 42.2 ± 13.1 |  | 42.2 ± 13.1 |  | 39.2 ± 10.4 |  | < 0.001 |
| **Age group (years)** |  |  |  |  |  |  |  |
| 20-49 | 73,069 | (73.1) | 72,888 | (73.1) | 181 | (86.2) | < 0.001 |
| 50-59 | 16,755 | (16.8) | 16,735 | (16.8) | 20 | (9.5) |  |
| 60-69 | 6,644 | (6.6) | 6,638 | (6.7) | 6 | (2.9) |  |
| 70+ | 3,525 | (3.5) | 3,522 | (3.5) | 3 | (1.4) |  |
| **Occupation** |  |  |  |  |  |  |  |
| Housewife | 10,685 | (10.7) | 10,679 | (10.7) | 6 | (2.9) | < 0.001 |
| Healthcare worker | 10,183 | (10.2) | 10,151 | (10.2) | 32 | (15.2) |  |
| Other healthcare-related | 22,303 | (22.3) | 22,195 | (22.2) | 108 | (51.4) |  |
| Student | 919 | (0.9) | 919 | (0.9) | 0 | (0) |  |
| Other | 55,903 | (55.9) | 55,839 | (56.0) | 64 | (30.5) |  |
| **Disease severity (at primary infection) <sup>a</sup>** |  |  |  |  |  |  |  |
| Mild - moderate | 81,018 | (81.0) | 80,827 | (81.0) | 191 | (91.0) | < 0.001 |
| Severe | 18,975 | (19.0) | 18,956 | (19.0) | 19 | (9.0) |  |
| *Personal history of:* |  |  |  |  |  |  |  |
| **Obesity (BMI 30 or higher)** |  |  |  |  |  |  |  |
| No | 81,531 | (81.5) | 81,353 | (81.5) | 178 | (84.8) | 0.228 |
| Yes | 18,462 | (18.5) | 18,430 | (18.5) | 32 | (15.2) |  |
| **Type 2 diabetes mellitus** |  |  |  |  |  |  |  |
| No | 86,909 | (86.9) | 86,718 | (86.9) | 191 | (91.0) | 0.082 |
| Yes | 13,084 | (13.1) | 13,065 | (13.1) | 19 | (9.0) |  |
| **Arterial hypertension** |  |  |  |  |  |  |  |
| No | 82,167 | (82.2) | 81,991 | (82.2) | 176 | (83.8) | 0.535 |
| Yes | 17,826 | (17.8) | 17,792 | (17.8) | 34 | (16.2) |  |
| **Immunosuppression <sup>b</sup>** |  |  |  |  |  |  |  |
| No | 98,830 | (98.8) | 98,627 | (98.8) | 203 | (96.7) | 0.003 |
| Yes | 1,163 | (1.2) | 1,156 | (1.2) | 7 | (3.3) |  |
| **Chronic kidney disease** |  |  |  |  |  |  |  |
| No | 98,440 | (98.5) | 98,239 | (98.5) | 201 | (95.7) | 0.001 |
| Yes | 1,553 | (1.5) | 1,544 | (1.5) | 9 | (4.3) |  |
| **Chronic obstructive pulmonary disease** |  |  |  |  |  |  |  |
| No | 98,872 | (98.9) | 98,667 | (98.9) | 205 | (97.6) | 0.083 |
| Yes | 1,121 | (1.1) | 1,116 | (1.1) | 5 | (2.4) |  |
| **Asthma** |  |  |  |  |  |  |  |
| No | 96,906 | (96.9) | 96,705 | (96.9) | 201 | (95.7) | 0.315 |
| Yes | 3,087 | (3.1) | 3,078 | (3.1) | 9 | (4.3) |  |
| **Cancer (any site)** |  |  |  |  |  |  |  |
| No | 99,744 | (99.7) | 99,535 | (99.7) | 209 | (99.5) | 0.508 |
| Yes | 249 | (0.3) | 248 | (0.3) | 1 | (0.5) |  |

Abbreviations: **SARS-COV-2**, Severe acute respiratory coronavirus 2; **SD**, Standard deviation; **BMI**, Body mass index.
Notes: 1) The absolute and relative (%) frequencies are presented, except if the mean is specified; 1) *p*-value from chi-square or t-tests are presented as corresponding.
<sup>a</sup> Severe illness included the register of dyspnea requiring hospital admission.
<sup>b</sup> Immunosuppression referred to any cause of the related deficiency except for type 2 diabetes mellitus or renal impairment.

In multiple analyses (Table 2), increasing age was associated with a reduced risk of reinfection (RR_*per year*_= 0.99997, 95% CI 0.99814-0.99958), as well as those with severe primary illness (RR= 0.9989, 95% CI 0.9981-0.9997). Compared with housewives, healthcare workers (RR= 1.0042, 95% CI 1.0030-1.0055) and other healthcare-related employees (RR= 1.0025, 95% 1.0012 -1.0039) showed an increased reinfection risk. Other high-risk conditions included the personal history of immunosuppression (RR= 1.0038, 95% 1.0011 - 1.0065) or chronic kidney disease (RR= 1.0039, 95% CI 1.0016-1.0063).

**Table 2.** Predictors of symptomatic laboratory-confirmed SARS-COV-2 reinfection, Mexico 2020

|  | **RR (95% CI), *p*** |  |  |  |  |  |
| --- | --- | --- | --- | --- | --- | --- |
|  | **Bivariate analysis** |  |  | **Multiple analysis** |  |  |
| **Gender** |  |  |  |  |  |  |
| Female | 1.0000 |  |  | 1.0000 |  |  |
| Male | 0.9998 | (0.9993 - 1.0004) | 0.574 | 1.0004 | (0.9997 - 1.0009) | 0.256 |
| **Age group (years)** |  |  |  |  |  |  |
| 20-49 | 1.0000 |  |  | 1.0000 |  |  |
| 50-59 | 0.9987 | (0.9980 - 0.9995) | 0.001 | 0.9989 | (0.9981 - 0.9997) | 0.006 |
| 60-69 | 0.9984 | (0.9973 - 0.9996) | 0.007 | 0.9984 | (0.9972 - 0.9996) | 0.009 |
| 70+ | 0.9983 | (0.9968 - 0.9999) | 0.039 | 0.9982 | (0.9966 - 0.9999) | 0.032 |
| **Occupation** |  |  |  |  |  |  |
| Housewife | 1.0000 |  |  | 1.0000 |  |  |
| Healthcare worker | 1.0043 | (1.0032 - 1.0054) | < 0.001 | 1.0042 | (1.0030 - 1.0055) | < 0.001 |
| Other healthcare-related | 1.0026 | (1.0013 - 1.0038) | < 0.001 | 1.0025 | (1.0012 - 1.0039) | < 0.001 |
| Student | 0.9994 | (0.9964 - 1.0025) | 0.721 | 0.9993 | (0.9961 - 1.0024) | 0.646 |
| Other | 1.0006 | (0.9996 - 1.0015) | 0.227 | 1.0005 | (0.9994 - 1.0016) | 0.337 |
| **Disease severity (at primary infection) <sup>a</sup>** |  |  |  |  |  |  |
| Mild - moderate | 1.0000 |  |  | 1.0000 |  |  |
| Severe | 0.9987 | (0.9979 - 0.9994) | < 0.001 | 0.9989 | (0.9981 - 0.9997) | 0.007 |
| *Personal history of:* |  |  |  |  |  |  |
| **Obesity (BMI 30 or higher)** |  |  |  |  |  |  |
| No | 1.0000 |  |  | 1.0000 |  |  |
| Yes | 0.9996 | (0.9988 - 1.0003) | 0.228 | 0.9997 | (0.9989 - 1.0004) | 0.360 |
| **Type 2 diabetes mellitus** |  |  |  |  |  |  |
| No | 1.0000 |  |  | 1.0000 |  |  |
| Yes | 0.9993 | (0.9984 - 1.0001) | 0.082 | 0.9996 | (0.9987 - 1.0005) | 0.333 |
| **Immunosuppression <sup>b</sup>** |  |  |  |  |  |  |
| No | 1.0000 |  |  | 1.0000 |  |  |
| Yes | 1.0040 | (1.0013 - 1.0066) | 0.003 | 1.0038 | (1.0011 - 1.0065) | 0.005 |
| **Chronic kidney disease** |  |  |  |  |  |  |
| No | 1.0000 |  |  | 1.0000 |  |  |
| Yes | 1.0038 | (1.0015 - 1.0061) | 0.001 | 1.0039 | (1.0016 - 1.0063) | 0.001 |

Abbreviations: **RR**, Risk ratio; **CI**, Confidence interval; **BMI**, Body mass index.
Notes: 1) Generalized linear regression models were used to obtain RR and 95% CI; 2) Multiple regression coefficients were adjusted by variables listed in the table.
<sup>a</sup> Severe illness included the register of dyspnea requiring hospital admission.<sup>b</sup> Immunosuppression referred to any cause of the related deficiency except for type 2 diabetes mellitus or renal impairment.

## Discussion

Our results suggest that symptomatic SARS-COV-2 reinfection is a rare phenomenon and factors associated with its risk were characterized. However, these results must be carefully considered since currently there is not a well-defined criterion for SARS-COV-2 reinfection [1]. All enrolled subjects reported disappearance of symptoms from primary infection. The used cutout point to identify potential cases of reinfection seemed to be epidemiologically useful since is according to the observed IgG antibodies titers decay in recovered COVID-19 patients [6].

According to our findings, healthcare workers and other related employees (e.g., medical assistants, dentists, etc.) are at increased risk of SARS-COV-2 symptomatic reinfections, which sounds plausible given the increased risk of exposure among these subjects [7]. Mild COVID-19 patients at primary episodes may also be at greater risk of reinfection, which may be secondary to lower antibody titers compared with pneumonia patients [8]. The association between immunosuppression and [2] renal impairment with COVID-19 risk has been widely discussed [4, 9]. If later replicated, further research is needed to identify factors determining a decreased reinfection risk among older participants and after adjusting by multiple exposures. We hypothesize that a reduced COVID-19 awareness among younger subjects may be implied, at least partially.

## Conclusions

To the best of our knowledge, this is the first study evaluating predictors of symptomatic SARS-COV-2 reinfection in a large subset of individuals and populations at high-risk were identified. Clinical and epidemiological research regarding SARS-COV-2 reinfection has immediate implications for public health policies focusing on reducing viral spread, including vaccine-related and social-distancing interventions.

## Data Availability

All data generated or analyzed during this study are included in this published article.

## Funding

This study was self-funded by the researchers.

## Ethical approval

The Health Research Committee 601 of the Mexican Institute of Social Security provided approval (R-2020-601-015).

## Conflict of interest

None to declare.

## Notes

### Competing Interest Statement

The authors have declared no competing interest.

